# Social Determinants of Health and GLP-1 RA Use Among Patients with Type 2 Diabetes and Stage 2 CKM: Insights from NHANES 2005–2020

**DOI:** 10.64898/2026.08.18.26360763

**Authors:** Qi Jian, Mark Segal, Hui Shao, Naykky Ospina, Tianze Jiao

**Affiliations:** Department of Pharmaceutical Outcomes and Policy, College of Pharmacy, University of Florida, Gainesville; Center for Drug Evaluation and Safety, University of Florida, Gainesville; Division of Nephrology, Hypertension & Renal Transplantation, Department of Medicine, College of Medicine, University of Florida, Gainesville, Florida; Hubert Department of Global Health, Emory Rollins School of Public Health, Emory University, Atlanta, GA; Division of Endocrinology, Department of Medicine, University of Florida, Gainesville

**Author notes:** **Corresponding Author:** Tianze Jiao, PhD Assistant Professor, Department of Pharmaceutical Outcomes & Policy College of Pharmacy, University of Florida, 1889 Museum Road, Rm 6301, Gainesville FL 32611.

## Abstract

**Background:** Cardiovascular-Kidney-Metabolic (CKM) syndrome encompasses interconnected conditions such as type 2 diabetes (T2D), hypertension, hypertriglyceridemia, metabolic syndrome (MetS), and chronic kidney disease (CKD). As CKM progresses, cardiorenal risks increase. Although Glucagon-like peptide-1 receptor agonists (GLP-1 RA) have demonstrated cardiorenal and cardiometabolic benefits, offering an opportunity to slow CKM progression, their use may vary across social determinants of health (SDoH) and stage 2 CKM subgroups.

**Objective:** To evaluate the influence of SDoH on access to GLP-1 RA among patients with T2D and other stage 2 CKM conditions.

**Methods:** This cross-sectional study used data from the U.S. National Health and Nutrition Examination Survey (NHANES), 2005–2020. Adults aged ≥30 years with T2D and/or other stage 2 CKM conditions were included. Weighted descriptive analysis, multivariable logistic regression and LASSO were applied to assess associations between SDoH and GLP-1 RA use.

**Results:** Among 4,520 participants (representing approximately 84.0 million U.S. adults), weighted mean age was 61.4 years, 48.9% were female, and 61.5% were non-Hispanic White. Among participants with T2D, GLP-1 RA use was higher among individuals with higher education (3.39% vs 1.43%), private insurance (3.00% vs 0.58%), and higher income (4.70% vs 1.87%), while no use was observed among those without routine places for care. In adjusted analyses, individuals with lower income, less than high school education, lack of insurance, and being unmarried had 64%, 51%, 81%, and 40% lower likelihood of GLP-1 RA use, respectively. LASSO identified income, education, insurance, and access to care as predictors. Lower income, lower educational attainment, and lack of insurance were associated with 48%, 34%, and 79% lower likelihood of GLP-1 RA use, respectively, adjusting for age, sex, and race/ethnicity.

**Conclusion:** SDoH-driven disparities limit GLP-1 RA access. Expanding GLP-1 RA access by addressing socioeconomic barriers is critical to slowing CKM progression, reducing cardiovascular risk, and mitigating health disparities.

## Background

Cardiovascular-Kidney-Metabolic (CKM) syndrome is a recently defined clinical condition encompassing the interrelated epidemics of metabolic disease, chronic kidney disease (CKD), and cardiovascular disease (CVD), which share overlapping pathophysiologic mechanisms and frequently coexist, resulting in progressive multi-organ dysfunction and elevated cardiovascular risk.^1^ To optimize monitoring and treatment for these patients, the American Heart Association introduced a CKM staging system (stages 0–4), among which stage 2 represents a critical inflection point, defined by the presence of metabolic risk factors, including type 2 diabetes (T2D), hypertension, hypertriglyceridemia, and moderate to high risk CKD, prior to the development of overt CVD.^2^ Strikingly, nearly 50% of U.S. adults meet criteria for stage 2 CKM, placing tens of millions at heightened risk for downstream cardiovascular and renal events.^2^ As advancing CKM stage is associated with markedly increased mortality risk,^3^ stage 2 CKM is emphasized as a key window for intervention to address risk factors and prevent progression to heart failure, atherosclerotic events, and kidney failure.^1^ Within this stage, T2D functions as a pivotal “gateway” condition, conferring a 2-to 4-fold higher risk of myocardial infarction and stroke, up to an 8-fold higher risk of heart failure, and accounting for approximately 40% of incident kidney failure through diabetic nephropathy.^4^ In effect, T2D accelerates the progression of CKM syndrome, propelling patients toward stage 3–4 events (subclinical and clinical CVD).^4^ This underpins intense interest in T2D as a key therapeutic target within stage 2 CKM.

In recent years, glucagon-like peptide-1 receptor agonists (GLP-1 RAs) have emerged as a breakthrough treatment for T2D. Beyond improving glycemic control through glucose-dependent insulin secretion,^5^ GLP-1 RAs promote clinically meaningful weight loss, directly counteracting the insulin resistance and dysfunctional adiposity central to CKM progression.^6^ Large cardiovascular outcome trials (e.g., LEADER, SUSTAIN-6, REWIND)^7,8,9^ demonstrate consistent reductions in major adverse cardiovascular events (MACE), with meta-analytic evidence showing a 14% reduction, alongside lower risks of cardiovascular mortality (13%), stroke (16%), and all-cause mortality (12%) compared to placebo.^10^ GLP-1 RAs also confer renal protection, reducing composite kidney outcomes by 17%, largely through marked reductions in albuminuria.^10^ Collectively, these effects position GLP-1 RAs as a compelling therapeutic strategy to modify the trajectory of stage 2 CKM before irreversible cardiovascular or renal damage occurs. Reflecting this broad therapeutic potential, an estimated 136.8 million U.S. adults are clinically eligible for semaglutide across indications including T2D, obesity, and cardiovascular risk reduction,^11^ driven by the high prevalence of cardiometabolic conditions such as hypertension, dyslipidemia, metabolic syndrome (MetS), and moderate CKD.^12^ Despite this vast eligible population, real-world utilization of GLP-1 RAs, particularly among individuals within stage 2 CKM, remains poorly characterized, leaving the magnitude of unmet therapeutic need and treatment gaps largely unknown.

Social determinants of health (SDoH) may suppress the real-world utilization of these already underused therapies on patients with T2D or obesity. Prior studies have shown low overall uptake of GLP-1 RAs in the United States: in a cohort of over 1.1 million adults T2D, fewer than 10% initiated GLP-1 RA, with substantially lower use among Black and Hispanic individuals and those with lower income, even after accounting for clinical factors.^13^ Similarly, among U.S. adults with obesity, only 2.3% received a GLP-1 RA, with markedly lower use in rural communities and groups with lower socioeconomic status.^14^ Restrictive coverage policies, including limited Medicaid coverage for obesity indications, prior authorization requirements, and high out-of-pocket costs, further constrain access.^15^ As a result, many patients who may benefit from GLP-1 RA remain untreated. This disconnection between therapeutic efficacy and limited treatment access exposes a critical evidence gap. Although CKM has been recognized as a unified cardiometabolic framework, population-level evidence remains limited in capturing patients with stage 2 CKM, and the association between SDoH and GLP-1 RA utilization among this population remains understudied.^13,14^ Thus, the objective of this study is to quantify real-world utilization of GLP-1 RAs among U.S. adults with stage 2 CKM and to examine how utilization varies across key SDoH strata and stage 2 CKM disease profile, generating evidence to inform clinical strategies aimed at ensuring appropriate cardiometabolic care across all patient subgroups.

## Methods

### Data Source and population

We conducted a cross-sectional analysis utilizing data from the National Health and Nutrition Examination Survey (NHANES), covering seven cycles from 2005 to March 2020. Data collection for the 2019–2020 cycle was incomplete due to disruptions from the COVID-19 pandemic. Thus, data collected through March 2020 were combined with the 2017–2018 cycle to maintain national representativeness.

The analytic sample included adults aged 30 years and older with type 2 diabetes, either alone or in combination with additional conditions of stage 2 CKM. Stage 2 CKM was defined following guidelines recommendations,^1,2^ including hypertension (blood pressure ≥140/90 mmHg or antihypertensive medication use), hypertriglyceridemia (fasting triglycerides ≥135 mg/dL), MetS (defined by NCEP ATP III criteria), and moderate-to high-risk chronic kidney disease (CKD, defined using modified KDIGO eGFR and ACR risk categories), with detailed operational definitions provided in **Table S1**. Participants could meet criteria for multiple stage 2 CKM conditions simultaneously. Additionally, we conducted exploratory analyses to evaluate whether diagnoses of hypertension, obesity, polyendocrine metabolic ovarian syndrome (PMOS) and metabolic dysfunction-associated steatotic liver disease (MASLD) could serve as proxies for MetS, thereby avoiding the need to use biomarker-based criteria.^16,17,18^ Detailed definitions are provided in **Table S2**. Individuals were excluded if they had a diagnosis of type 1 diabetes or if they were missing on key variables required to operationalize conditions of stage 2 CKM, including biomarker-based measures (e.g., eGFR for CKD risk classification).

### Outcome

The primary outcome was current use of a GLP-1 RA, identified through self-reported prescription medication use collected during the NHANES survey interview, including the use of specific agents such as exenatide, liraglutide, dulaglutide, semaglutide, lixisenatide, or albiglutide.

### Exposure

Exposure variables included SDoH mapped to the Healthy People 2030 framework. These included 5 domains: (1) economic stability (employment status [employed/not employed], household income-to-poverty ratio [high income: ≥300%/low income: <300%], and food insecurity [full security/marginal or low security]), (2) education access and quality (education level [high school or above/less than high school]), (3) healthcare access and quality (insurance coverage [insured/not insured] and usual source of care [has routine place for healthcare/no routine place for healthcare]), (4) neighborhood and built environment (housing status [own home/rent or other arrangement]), and (5) social and community context (marital status [married/not married]).^19,20^ Neighborhood and built environment indicators, such as housing stability, were only available for NHANES 2005–2016 and were not collected in the 2017–2020 cycle; therefore, these variables were only included for the years in which data were available.

### Statistical analysis

Descriptive analyses were first conducted to estimate the proportion of GLP-1 RA use across categories of SDoH, stratified by combinations of CKM conditions. These proportions were then visualized using a two-dimensional heatmap to illustrate disparities in GLP-1 RA use across SDoH strata and CKM subgroups. To improve contrast and reduce the influence of extreme values, heatmap values were square root-transformed for coloring. Additionally, to assess associations between individual SDoH indicators and the likelihood of GLP-1 RA use, we conducted separate multivariable logistic regression models for individual SDoH domain. These models estimated adjusted odds ratios (ORs) and 95% confidence intervals (CIs), controlling for age, sex, and race/ethnicity. To select the top influential SDoH predictors, we applied Least Absolute Shrinkage and Selection Operator (LASSO) logistic regression with 3-fold cross-validation. All statistical analyses were conducted using SAS OnDemand for Academics and RStudio (version 2024.09.1+394). A two-sided p-value of < 0.05 was considered statistically significant.

## Results

As shown in Figure 1, after excluding individuals with missing key covariates and restricting the analytic sample to adults aged ≥30 years with T2D, a total of 4,520 participants were included in the unweighted sample, corresponding to a nationally representative weighted population of 84,028,675 U.S. adults.

**Figure 1.**
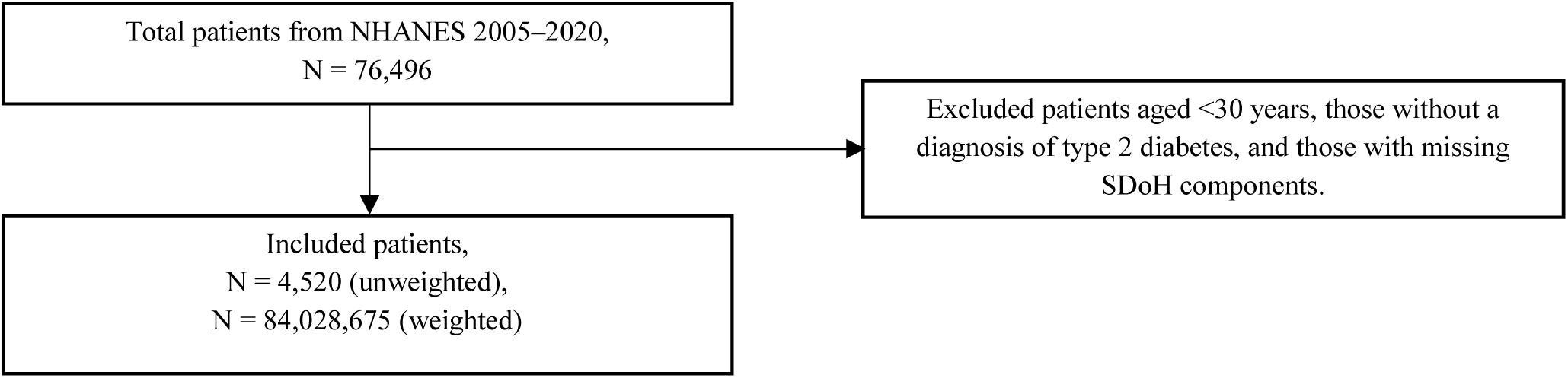
Flowchart.

The weighted mean age was 61.38 years (SE: 0.26), and 48.87% were female. The racial/ethnic distribution was 61.51%, 15.85% and 22.64% for non-Hispanic White, non-Hispanic Black, and other race/ethnicity, respectively (Table 1). Among adults in our cohort, cardiometabolic comorbidities were common: 73.85%, 79.51%, 23.46%, and 21.42% of them were diagnosed with hypertension, MetS, hypertriglyceridemia and moderate-to-high risk CKD, respectively. In exploratory analyses, 23.86% had MASLD, and 10.24% had PMOS (only available 2011–2020 cycles). Across SDoH domains, 59.10% had a poverty-income ratio <300%, 26.29% reported marginal or low food security, and 25.18% had less than high school education. Within the healthcare access domain, 90.64% were insured and 96.74% reported a routine place for care. In the neighborhood and social context domains, 26.99% lived in rented or other non-owned housing (available 2005–2016 cycles), and 40.19% were not married.

**Table 1.** Baseline characteristics.

|  | All (unweighted)<br>(n=4,520) | All (weighted)<br>(N= 84,028,675) |
| --- | --- | --- |
| **Age, years** |  |  |
| Mean (SE) | 63.30<br>(0.17) | 61.38<br>(0.26) |
| <65 | 2,331<br>(51.57%) | 48,637,998<br>(57.88%) |
| ≥65 | 2,189<br>(48.43%) | 35,390,677<br>(42.12%) |
| **Female gender** | 2,131<br>(47.15%) | 41,064,504<br>(48.87%) |
| **Race** |  |  |
| White | 1,588<br>(35.13%) | 51,681,939<br>(61.51%) |
| Black | 1,274<br>(28.19%) | 13,320,373<br>(15.85%) |
| Other | 1658<br>(36.68%) | 19,026,362<br>(22.64%) |
| **Medical status** |  |  |
| Hypertension | 3,344<br>(73.98%) | 62,053,637<br>(73.85%) |
| Metabolic syndrome | 3,426<br>(75.80%) | 66,807,880<br>(79.51%) |
| Hypertriglyceridemia | 871<br>(19.27%) | 19,716,600<br>(23.46%) |
| Moderate to high-risk chronic kidney disease (CKD) | 1,013<br>(22.41%) | 18,001,303<br>(21.42%) |
| Polyendocrine Metabolic Ovarian Syndrome (PMOS)* | 291<br>(10.02%) | 5,100,151<br>(10.24%) |
| Metabolic Dysfunction-Associated Steatotic Liver Disease (MASLD) | 1,000<br>(22.12%) | 1,108,015<br>(23.86%) |
| **SDoH** |  |  |
| **Economic stability** |  |  |
| **Employment status** |  |  |
| Employed | 1,797<br>(39.76%) | 23,616,204<br>(28.10%) |
| Unemployed | 2,723<br>(60.24%) | 60,412,470<br>(71.90%) |
| **Poverty income ratio** |  |  |
| <300 | 3158<br>(69.87%) | 49,664,599<br>(59.10%) |
| ≥300 | 1,362<br>(30.13%) | 34,364,075<br>(40.90%) |
| **Food security status** |  |  |
| Full security | 2,970<br>(65.71%) | 61,937,952<br>(73.71%) |
| Marginal, low, or very low security | 1,550<br>(34.29%) | 22,090,723<br>(26.29%) |
| **Education access and quality** |  |  |
| Less than high school | 1,542<br>(34.12%) | 21,160,323<br>(25.18%) |
| High school or above | 2,978<br>(65.88%) | 62,868,352<br>(74.82%) |
| **Healthcare access and quality** |  |  |
| **Insurance status** |  |  |
| Insured | 4003<br>(88.56%) | 76,165,033<br>(90.64%) |
| Not insured | 517<br>(11.44%) | 7,863,642<br>(9.36%) |
| **Access to healthcare** |  |  |
| Routine place to go to healthcare | 4344<br>(96.11%) | 81,285,905<br>(96.74%) |
| No routine place to go to healthcare | 176<br>(3.89%) | 2,742,769<br>(3.26%) |
| **Neighborhood and built community** |  |  |
| **House status<sup>†</sup>** |  |  |
| Own home | 2,294<br>(66.88%) | 57,884,382<br>(73.01%) |
| Rent or other arrangement | 1,136<br>(33.12%) | 21,403,159<br>(26.99%) |
| **Social and community context** |  |  |
| **Marital status** |  |  |
| Married | 2,540<br>(56.19%) | 50,259,501<br>(59.81%) |
| Not married | 1980<br>(43.81%) | 33,769,173<br>(40.19%) |

Notes: \*PMOS data is only available from 2011 to 2020.
†House status data is only available from 2005-2016.

GLP-1 RA use varied across combinations of type 2 diabetes and additional stage 2 CKM components, as well as across SDoH strata (Figure 2). All reported percentages represent the proportion of individuals using GLP-1 RA within each specific disease and SDoH combination subgroup. Among individuals with T2D alone, GLP-1 RA use was low across subgroups, with use up to 4.70%: GLP-1 RA use was more common among those with higher educational attainment (3.39% vs 1.43%), insurance coverage (3.00% vs 0.58%), and higher income (4.70% vs 1.87%), whereas no use was observed among individuals without a routine place for care.

**Figure 2.**
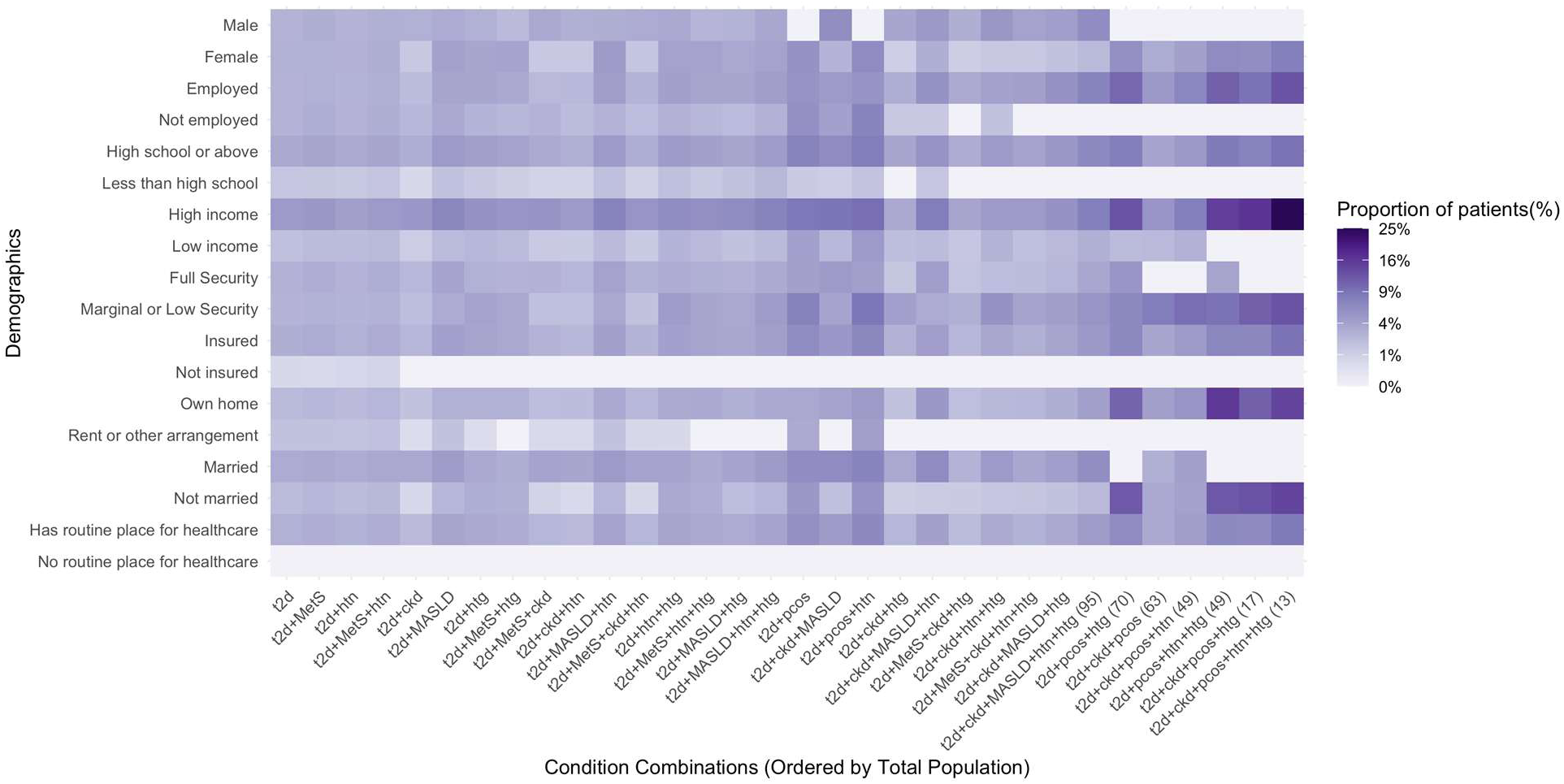
Heatmap of GLP-1 RA Utilization Across Social Determinants of Health and CKM Stage 2 Subgroups*†. *Abbreviations: t2d: type 2 diabetes; htn: hypertension; htg: hypertriglyceridemia; ckd: chronic kidney disease; MASLD: metabolic dysfunction-associated steatotic liver disease; pmos: polyendocrine metabolic ovarian syndrome; MetS: metabolic syndrome. High income: poverty-income ratio ≥300%; Low income: poverty-income ratio <300%. Full Security: full food security; Marginal or Low Security: marginal, low, or very low food security. Has routine place for healthcare: reported having a usual source of healthcare; No routine place for healthcare: reported no usual source of healthcare. †The color intensity of each tile represents the weighted proportion (%) of GLP-1 RA users within each subgroup. To improve visualization and contrast across groups with small absolute differences, proportions were square-root transformed for the heatmap color scaling. However, the legend labels display the original untransformed weighted percentages for interpretability.

Across many demographic and SDoH strata, GLP-1 RA use was more common in subgroups with greater CKM burden compared with T2D alone. For example, among males, use increased from 2.64% in T2D alone to 5.00% in T2D with CKD, hypertension, and hypertriglyceridemia; similar patterns were observed across education, marital status, and food security strata.

Substantial disparities were observed across socioeconomic and access-related factors, with the most pronounced differences evident in higher-burden CKM combinations. In comparable disease strata, GLP-1 RA use was markedly higher among individuals with at least a high school education compared with those without (e.g., 4.72% vs 0.00% in T2D with CKD, hypertension, and hypertriglyceridemia), and among those with insurance compared with those without (e.g., 3.57% vs 0.00% in T2D with CKD, hypertension, and hypertriglyceridemia; 4.05% vs 0.00% in T2D with hypertension and hypertriglyceridemia). Income-related differences were also evident, with higher use among individuals with high income compared with those with low income (e.g., 4.55% vs 2.68% in T2D with CKD, hypertension, and hypertriglyceridemia). Notably, individuals without a routine place for healthcare had no observed GLP-1 RA use across any disease combination. All above reported percentages reflect original weighted proportions, and square-root transformation was applied solely for visualization.

In multivariable logistic regression models adjusted for age, sex, and race/ethnicity (Figure 3), several SDoH indicators were independently associated with GLP-1 RA use. Lower income (<300% poverty-income ratio) was associated with substantially lower odds of GLP-1 RA use (OR 0.36, 95% CI 0.24–0.55). Individuals with less than a high school education had 51% lower odds of treatment compared with those with higher education (OR 0.49, 95% CI 0.30–0.79).

**Figure 3.**
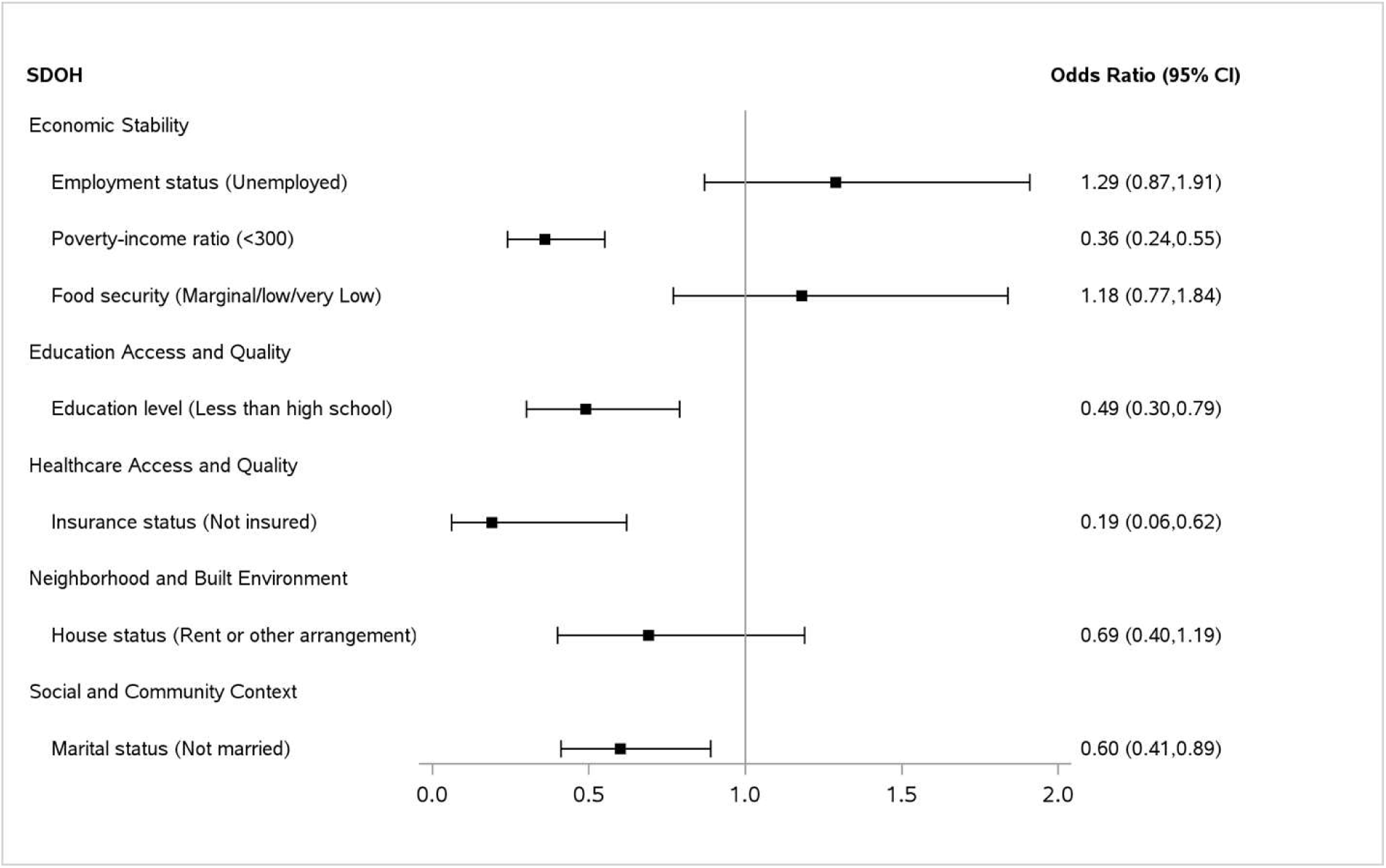
Adjusted Odds Ratios for GLP-1 Receptor Agonist Use by Social Determinants of Health, Adjusted for Age, Sex, and Race/Ethnicity.

Uninsured individuals demonstrated the strongest association, with markedly reduced odds of GLP-1 RA use (OR 0.19, 95% CI 0.06–0.62), corresponding to an 81% lower likelihood of treatment. Not being married was associated with lower utilization (OR 0.60, 95% CI 0.41–0.89). Employment status, food insecurity, and housing status were not significantly associated with GLP-1 RA use.

In LASSO model, the optimal penalty parameter (λ) was selected based on the 1-standard-error rule (Figure S1). Under this criterion, four SDoH variables were retained with non-zero coefficients: poverty–income ratio, education level, insurance status, and routine access to healthcare. Although no GLP-1 RA use was observed among participants without a routine place for care, the LASSO penalty constrained the coefficient estimate to remain finite, allowing routine access to healthcare to be retained as a strong predictor. However, this complete separation caused the coefficient estimate to diverge in the subsequent unpenalized logistic regression, resulting in model non-convergence. Therefore, routine access to healthcare was excluded from the adjusted model, while the other three SDoH variables were included along with age group, sex, and race. In the adjusted analysis (Table 2), lower income, and lack of insurance were associated with 48%, and 79% lower odds of GLP-1 RA use, respectively (OR = 0.52, 95% CI: 0.35–0.75; OR = 0.21, 95% CI: 0.05–0.58), yet the association was statistically insignificant for lower educational attainment (OR = 0.66, 95% CI: 0.40–1.07). Older age (≥65 years) was also associated with 55% lower odds of use (OR = 0.45, 95% CI: 0.30–0.66), while non-Hispanic White individuals had higher odds of GLP-1 RA use compared with the “Other” group (OR = 1.87, 95% CI: 1.17–3.04, p = 0.010).

**Table 2.** Adjusted Association of LASSO-Selected Social Determinants of Health with GLP-1 Receptor Agonist Use.

| Variable | OR (95% CI) | p-value |
| --- | --- | --- |
| Poverty-income ratio <300% | 0.52 (0.35–0.75) | <0.001 |
| Less than high school education | 0.66 (0.40–1.07) | 0.10 |
| Uninsured | 0.21 (0.05–0.58) | 0.01 |
| Age ≥65 years | 0.45 (0.30–0.66) | <0.001 |
| Female (vs male) | 1.16 (0.81–1.67) | 0.42 |
| Non-Hispanic Black (vs Other) | 1.58 (0.97–2.62) | 0.07 |
| Non-Hispanic White (vs Other) | 1.87 (1.17–3.04) | 0.01 |

## Discussion

### Key finding and population level number about unmet need

In this nationally representative NHANES study of patients with T2D and an additional stage 2 CKM syndrome, we found that use of GLP-1 RAs remained strikingly low in many high-risk patient groups and varied across SDoH subgroups. Lower GLP-1 RA use was consistently observed among individuals with lower income, lower education, and no insurance coverage. Its uptake was also low in patients with T2D plus additional comorbidity of stage 2 CKM, even though modern guidelines recommend GLP-1 RAs for most T2D patients with CKD or cardiovascular risk.^21,22^ Translating our results to the U.S. population highlights the substantial scale of this unmet need. An estimated 32.5 million U.S. adults with T2D and CKD or hypertriglyceridemia are not receiving GLP-1 RAs, the group for whom guidelines most strongly recommended these agents due to the greatest cardiorenal benefits. Similarly, approximately 4.5 million diabetic individuals with comorbid CKD and MASLD are not using GLP-1 RAs, representing missed opportunities for renal and cardioprotective benefits. From another perspective, these differences became even more apparent when examined across SDoH-defined population subgroups. Roughly 23.0 million unemployed, 48.9 million low-income, and 7.8 million uninsured Americans with T2D who could benefit from GLP-1 RAs are not accessing them. Together, these findings suggest that low GLP-1 RA utilization disproportionately affects population subgroups with both elevated CKM risk burden and less favorable SDoH characteristics. This underscores that the underuse of GLP-1 RAs is not a marginal issue, but a population-level gap, leaving millions of high-risk individuals undertreated.

### Why This Pattern Matters in Stage 2 CKM

By definition, stage 2 CKM means major metabolic risk factors are present – e.g. T2D, hypertension, dyslipidemia, or early CKD – yet overt cardiovascular disease has not yet occurred.^1^ It is the most prevalent CKM stage that nearly half of U.S. adults fall into,^2^ and is a critical window for intervention. The treatment goal in stage 2 is to prevent progression to stage 3 or 4 where actual organ damage and clinical CVD occur. GLP-1 RAs are uniquely suited to CKM stage 2, as they directly target the dominant pathogenic processes at this transitional stage. Clinically, GLP-1 RAs induce substantial weight loss and improve insulin sensitivity, mitigating the obesity–insulin resistance axis that underlies T2D and metabolic liver disease.^23,24,25^ Beyond metabolic control, GLP-1 RAs exert pleiotropic vascular effects, including reductions in systemic inflammation and improvements in endothelial function, mechanisms that are central to atherosclerotic disease progression and cardiovascular risk.^25,26^ GLP-1 RAs also exert renoprotective effects by reducing albuminuria, slowing eGFR decline, and lowering adverse kidney outcomes.^27^ Failure to initiate GLP-1 RAs during stage 2 CKM represents a critical missed opportunity for risk modification before the onset of irreversible cardiovascular or renal damage.

### SDoH gradient

Our analysis identified several SDoH factors as independent predictors of GLP-1 RA underuse – notably lower income, less education, lack of insurance, and no routine place for healthcare (selected by LASSO) among patients with stage 2 CKM. Our results align with and extend the prior literature on demonstrating variation in GLP-1 RA utilization among patients with T2D and across subgroups. Prior research has shown that overall GLP-1 RA uptake among patients with T2D has historically been low: among 1.18 million insured patients with T2D, only 7.7% were treated with a GLP-1 RAs from 2015 to 2019.^13^ Prior studies have quantified these disparities, reporting that individuals with some college education or higher had approximately 1.8–2.1 times higher odds of GLP-1 RA use, and those with higher income-to-poverty ratios had roughly twofold higher odds of treatment compared with lower-income groups.^28^ The high cost of GLP-1 RAs, often over $900 per month without insurance, puts them out of reach for uninsured or underinsured patients.^29^ The lack of a regular source of healthcare further widened this gap by reducing opportunities for in-person communication between patients and healthcare providers. Educational attainment plays a role as well: lower education may translate into less health literacy, and less awareness of newer treatment options.

### Hypertension, Obesity, PMOS and MASLD: Underappreciated Metabolic Risk Signals

One notable finding was that a proxy definition based on hypertension, obesity, PMOS, and MASLD demonstrated substantial overlap with MetS, capturing approximately 98% of MetS cases in our study. The PMOS and MASLD are well-known to confer elevated cardiometabolic risk,^30,31^ both conditions are closely linked to insulin resistance and broader cardiometabolic dysregulation,^32^ yet they are not included in the formal NCEP ATP III definition of MetS. Existing clinical evidence consistently supports the association between PMOS and adverse cardiometabolic profiles, underscoring the biological plausibility of shared metabolic risk.

Together with hypertension and obesity, PMOS and MASLD are clinically relevant indicators of elevated metabolic risk and may serve as proxies for MetS when biomarker data are unavailable.

### Strengths

Our study has several notable strengths. First, by leveraging NHANES data, we analyzed a nationally representative sample of U.S. adults, which enhances generalizability. Second, we applied the CKM staging framework to real-world data. By focusing on stage 2 CKM, we bring a novel, clinically relevant perspective – aligning our analysis with the idea of intervening early in the trajectory of CKM progression. This framing goes beyond a traditional study by explicitly incorporating multi-organ risk. Third, our study extends prior literature by simultaneously examining the interplay of multiple cardiometabolic comorbidities and SDoH within a single national representative database. Whereas previous studies often evaluated GLP-1 RA uptake along isolated dimensions, such as racial disparities or use among patients with established cardiovascular disease, we jointly considered a broad spectrum of clinical factors (including CKD, hypertension, hyperlipidemia, MASLD, and PMOS) alongside multiple SDoH indicators. This integrated framework allowed us to capture heterogeneity in treatment patterns that may be obscured when clinical or social factors are examined separately. Fourth, we employed LASSO regression for variable selection to efficiently evaluate a set of candidate predictors and identify the most robust factors associated with GLP-1 RA use while minimizing overfitting. This approach strengthened inference in a high-dimensional setting and provided a more nuanced understanding of GLP-1 RA uptake. Finally, we translated our prevalence findings into population-level impact, which is valuable for policy and public health discussions. By quantifying the number of people left undertreated, we highlight the significance of the issue in the U.S.

### Limitations

We acknowledge several important limitations. First, the NHANES design is cross-sectional, capturing a snapshot of GLP-1 RA use and associated factors at a single time point, which precludes causal inference and assessment of temporal relationships. Second, GLP-1 RA treatment status was ascertained using self-reported medication use, which is subject to recall bias and potential misclassification. Although over 80% of participants had medication containers verified,^33,34^ reducing the likelihood of substantial misclassification, some degree of measurement error may persist. Furthermore, participants with missing data on key biomarkers used to define comorbidities (e.g., HbA1c for T2D) were excluded from the analysis, which may result in underrepresentation of certain subpopulations and introduce selection bias. Finally, several subgroup analyses had small sample sizes, leading to unstable estimates. Subgroups with very low uptake (<1%) should be interpreted with caution due to sparse data.

### Conclusion

Our nationally representative NHANES analysis reveals substantial underuse of GLP-1 RAs among stage 2 CKM patients, particularly those with multiple cardiometabolic comorbidities and less favorable SDoH characteristics, despite strong evidence of benefit at this disease stage. Future research should focus on implementation strategies and approaches to improve the access to GLP-1 RAs in clinically high-risk populations and better translate cardiometabolic therapeutic advances into real-world risk reduction.

## Data Availability

The National Health and Nutrition Examination Survey (NHANES) data files and survey documentation used in this study are publicly available and can be accessed through the NHANES website at https://wwwn.cdc.gov/nchs/nhanes/.

## Acknowledgement

None.

